# Urban Mobility and Population Health in The Hague: A Multi-Scale Analysis

**DOI:** 10.64898/2026.06.23.26356068

**Authors:** Hielke Muizelaar, Marcel Haas, Rimke Vos, Ilonca Vaartjes, Ester de Jonge, Lampros Stergioulas, Jessica Kiefte-de Jong, Marco Spruit

## Abstract

Urban mobility may provide insight into population health by reflecting how residents connect to services, resources, and urban systems. However, mobility patterns are also shaped by contextual factors, including socioeconomic, environmental, behavioural, and demographic conditions, complicating interpretation of mobility– health associations, particularly when mobility and health are measured at different spatial scales.

This ecological cross-sectional study examined associations between mobile-phone-derived mobility and population health in The Hague, the Netherlands, from January to July 2019. Mobility was measured as mean outgoing distance across eight regions, while contextual and health indicators were primarily available for 44 neighbourhoods. Health outcomes included cardiometabolic medication use, polypharmacy, and syndemic-based disease burden. Using a residual-based adjustment framework, mobility and health outcomes were modelled separately using the same contextual domains, and residual correlations assessed whether mobility retained health-relevant variation beyond shared contextual conditions.

Outgoing mobility varied substantially across regions and was strongly patterned by contextual factors. At the regional level, higher-than-expected mobility tended to coincide with lower-than-expected disease burden after contextual adjustment (ρ = −0.26 to −0.48). In cross-scale neighbourhood analyses, however, residual correlations were considerably weaker (ρ = −0.11 to 0.10). Cluster-bootstrap 95% confidence intervals included zero for all outcomes, and sensitivity analyses yielded similar results.

These findings suggest that aggregated mobility is better interpreted as a spatially and contextually embedded indicator of urban connectivity and population composition than as a standalone indicator of population health. The contrast between regional and neighbourhood-level findings highlights how geographic units can influence interpretation of mobility–health relationships.

## 1. Introduction

Population health and resilience in urban environments are increasingly understood as the result of multiple environmental, social, and behavioural factors. Rather than acting independently, these exposures co-occur and interact across space, shaping health behaviours, disease clustering, multimorbidity, and inequalities within cities (Vlahov et al., 2007; Bai et al., 2012; Verheij, 1996; Ompad et al., 2007). Factors such as social needs, access to healthcare and transportation, socioeconomic conditions, environmental quality, and social connectivity jointly influence the development and progression of chronic diseases (Heller et al., 2021; Seraphim et al., 2025). This is particularly relevant for multimorbidity, where multiple conditions co-occur within individuals and populations, often reflecting shared underlying social and environmental determinants. Understanding urban health requires consideration of multiple determinants, but also attention to the geographic scales and spatial units at which these determinants and health outcomes are measured and linked.

Within this context, urban mobility, defined as the movement of people within and between urban areas; including the frequency, distance, and purpose of travel, offers a potential lens through which these interacting factors can be observed. Mobility is inherently spatial and connects residential areas to wider urban systems of employment, services, social interaction, and transport. However, because mobility patterns are embedded within broader social and environmental contexts, their relationship with health may be difficult to interpret in isolation. For example, a region with longer average outgoing mobility may appear healthier, but this pattern could reflect higher income, better access to transport, or a different age composition rather than mobility itself. Recent work has shown that neighbourhood environmental characteristics cluster into structured patterns, and that different environmental indicators can capture distinct dimensions of the urban environment (Smit et al., 2026).

Previous research has demonstrated relevance of mobility for public health, particularly in the context of infectious disease transmission, where population movement plays a key role in shaping epidemic dynamics (Wesolowski et al., 2016; Lu et al., 2026; Roy et al., 2025). Beyond infectious-disease applications, mobility may reflect access to services, employment, and social networks and thereby broader urban resilience (da Silva Sassaron et al., 2025; Van der Craats et al., 2025; Krathaus et al., 2026; Östh et al., 2018). In disease contexts, mobility may also reflect both health status and place-based constraints: cardiometabolic conditions can limit daily movement, while neighbourhood environments shape access to services and goods, opportunities for physical activity, and broader participation in society (Hidayati et al., 2021). Therefore, mobility has been recognised as a potential indicator of community health and population resilience, reflecting the ability of individuals and communities to adapt and recover from changing conditions (Östh et al., 2018). From a geographical perspective, however, mobility can also be understood as an expression of the functional organization of the city: movement patterns connect places across administrative boundaries and reflect the interaction between population composition, accessibility, land use, and urban opportunities.

In recent years, mobile phone mobility data and other digital sources have been used to examine behaviour patterns, access to urban facilities, and inequalities in neighbourhoods, suggesting that mobility reflects broader social and environmental conditions (Palmer et al., 2013; Xu et al., 2025; Liu et al., 2023a). In this study, we use the term contextual conditions to refer to the broader area-level characteristics that may shape both mobility and health, including socioeconomic position, demographic and population composition, housing, lifestyle-related indicators, access to facilities, social vulnerability, and physical environmental characteristics. Studies that combine mobility with contextual factors have shown that these jointly contribute to spatial variation in health outcomes (i.e. Liu et al., 2024; Yang et al., 2022; Gao & Kwan, 2026). These findings underline the importance of considering multiple contextual factors rather than examining individual environmental or socioeconomic determinants in isolation. However, it remains unclear whether aggregated mobility provides additional health-relevant variation beyond these co-occurring contextual conditions, or whether it mainly reflects same underlying structures. This question is complicated further when mobility and contextual health indicators are available at different spatial resolutions, for example when mobility is measured for extended mobility regions while socioeconomic and health indicators are available for smaller administrative neighbourhoods.

Against this background, understanding the association between mobility and health is particularly relevant to identify areas characterised by lower resilience and higher disease burden, supporting targeted prevention and policy interventions. While prior work examined mobility alongside socioeconomic and environmental determinants, less is known about whether mobility retains health-relevant information after accounting for measured contextual conditions. As a result, it remains unclear whether mobility retains an independent association with health beyond the conditions that shape it.

In this study, we examine the spatial relationship between population mobility, contextual conditions, and population health in The Hague, The Netherlands. Using aggregated mobile phone data available for eight mobility regions and neighbourhood-level indicators, we analyse how average outgoing mobility relates to indicators of disease burden, including cardiometabolic medication prescriptions, polypharmacy, and a syndemic-based disease burden measure. The contrast between eight functional mobility regions and finer administrative neighbourhoods provides an explicit multi-scale setting in which to examine both contextual confounding and the consequences of linking data across spatial supports.

To account for influence of co-occurring contextual factors, we adopt a residual-based modelling approach in which mobility is first modelled as a function of selected contextual domains, including demographic, socioeconomic, educational, migration-background, lifestyle, housing, facilities-access, physical environment, and social-vulnerability indicators. The remaining variation, representing mobility independent of these factors, is subsequently related to health outcomes. This approach allows us to assess whether deviations in mobility are associated with differences in population health.

In addition, we compare polypharmacy and a syndemic-based disease burden measure to evaluate whether the observed mobility-health association depends on how co-occurring disease burden is conceptualised. Polypharmacy captures the accumulation of medication use, whereas syndemics captures interacting conditions within their broader social and environmental context.

Overall, this study aims to examine how aggregated urban mobility relates to population health across functional and administrative spatial scales, and whether mobility retains an association with health beyond multiple measured contextual conditions. By combining a contextual adjustment framework with a multi-scale spatial analysis, we assess both the extent to which mobility captures health-relevant information beyond broader urban context and how this relationship depends on the spatial units through which mobility and health are represented.

## 2. Methods

This study is an ecological, cross-sectional multi-scale analysis conducted in The Hague, the Netherlands. The Hague is a densely urbanised coastal city in the western Netherlands and one of the country’s four largest cities, with approximately 538,000 residents at the start of 2019. The city is characterised by substantial socioeconomic and residential heterogeneity. Previous analyses by Statistics Netherlands showed marked neighbourhood differences in income, housing tenure, and perceived liveability, with relatively affluent neighbourhoods such as Vogelwijk and Benoordenhout contrasting with lower-income neighbourhoods such as Transvaal and Moerwijk (Van der Sangen, 2019). This combination of high urban density and pronounced neighbourhood-level variation makes The Hague a relevant setting for examining spatial relationships between mobility, contextual conditions, and population health.

### 2.1 Data Sources and Spatial Units

Mobility data were obtained from the Vodafone Mezuro dataset and consisted of daily aggregated movements between provider-defined Mezuro regions. These regions are the geographic units used by Mezuro to aggregate mobile-phone-derived origin–destination information. The nationwide zoning reflects the underlying mobile-network cell plan and the spatial accuracy attainable from call and data records; cities and villages generally constitute individual zones, while larger cities are subdivided into smaller urban areas (Wismans et al., 2018). For The Hague, the supplied dataset contained eight Mezuro regions.

VodafoneZiggo accounted for approximately 15-20% of mobile data usage in the Netherlands during the study period (Autoriteit Consument & Markt, 2019). The data were therefore interpreted as indicators of relative mobility differences rather than exact population-level movement estimates. As no information was available on how VodafoneZiggo users differed from the wider population, we could not determine whether this subpopulation was systematically biased with respect to characteristics relevant to mobility or health.

Mobility data covered January-July 2019, before the COVID-19 pandemic. A movement was defined as the presence of an individual in a Mezuro region other than their inferred home region, based on recurring location patterns; pass-through travel was excluded.

Contextual indicators were obtained from several sources. Statistics Netherlands (CBS) provided demographic and socioeconomic indicators, including population composition, income, housing characteristics, and accessibility of facilities, at municipality, district, neighbourhood, or postal-code level (Statistics Netherlands, 2019). Environmental indicators, including public green space and water coverage, were obtained from the Klimaateffectatlas (Stichting Climate Adaptation Services, 2026). Health-related contextual indicators were derived from the 2022 Health Monitor from the Dutch National Institute for Public Health and the Environment (RIVM, 2022), as the 2020 Health Monitor coincided with the COVID-19 pandemic. Neighbourhood estimates were derived using Small Area Estimation (Rao & Molina, 2015).

Health outcomes were obtained from the Extramural Leiden University Medical Center Academic Network (ELAN), a regional primary-care data infrastructure containing routinely recorded diagnoses and medication prescriptions (Kist et al., 2024; Ardesch et al., 2023). These data were aggregated into area-level indicators of cardiometabolic medication use, polypharmacy, and syndemic disease burden.

### 2.2 Spatial Alignment of Data

Mobility was analysed at its native spatial resolution of eight Mezuro regions, whereas contextual variables and health outcomes were primarily available at neighbourhood or postal-code level. The Mezuro regions therefore represent provider-defined mobility-analysis zones rather than administrative or empirically derived functional regions. Movements, including the outgoing-distance measure used in this study, were defined between these regions.

To link the datasets spatially, neighbourhoods were assigned to the Mezuro region with which they had the greatest geographic overlap. Where a neighbourhood intersected multiple Mezuro regions, the region containing the largest share of its area was selected. Overlap was generally high (median 98.6%, range 61.9-100%). Appendix A reports all assignments and corresponding overlap percentages.

This linkage introduces a change of spatial support between the mobility regions and administrative neighbourhoods. Figure 1 illustrates this structure: Panel A locates The Hague within the Netherlands, Panel B shows the eight Mezuro regions, Panel C shows the administrative neighbourhoods, and Panel D shows the overlap between both geographic systems. Neighbourhood numbers in Panel C correspond to Appendix A. Descriptive characteristics of the Mezuro regions are presented in Table 2.

**Figure 1.**
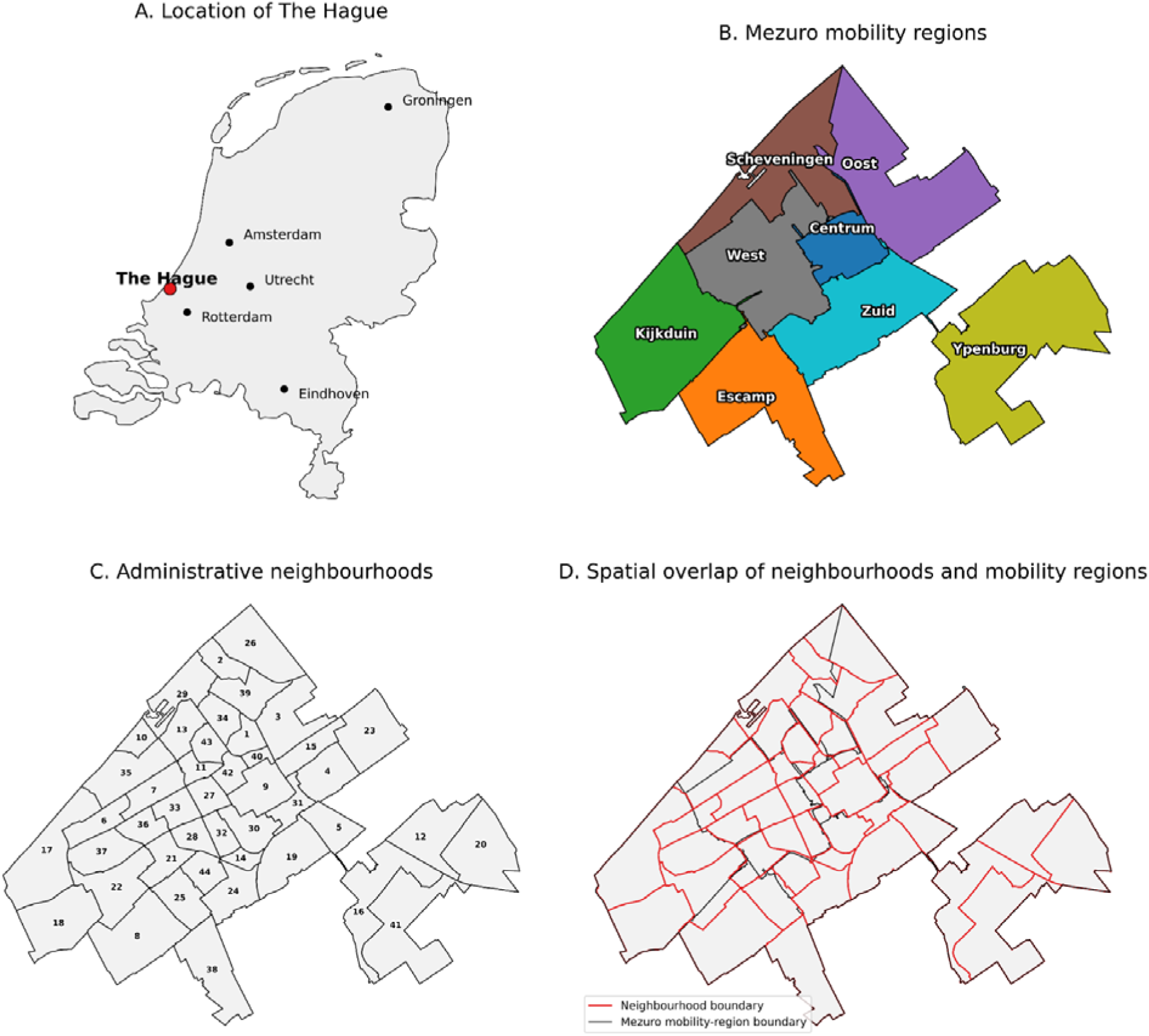
Study area and spatial units used in the analysis. (A) Location of The Hague within the Netherlands, with selected major Dutch cities shown for reference. (B) The eight Mezuro mobility regions used to represent aggregated mobility. (C) Administrative neighbourhoods in The Hague; numbers correspond to the neighbourhood-to-Mezuro assignments reported in Appendix A. (D) Administrative neighbourhood boundaries (red) overlaid on Mezuro mobility-region boundaries (black), illustrating the partial spatial mismatch between the two geographical systems.

### 2.3 Variables

#### 2.3.1 Mobility

Mobility was quantified primarily as the average outgoing distance per Mezuro region. For each Mezuro region and day, mean outgoing distance was calculated as a movement-volume-weighted average of centroid-to-centroid distances between origin–destination pairs. Distances to destination regions were weighted by the number of observed outgoing movements from the origin region to each destination. The resulting daily weighted means were then averaged across the study period to obtain one regional estimate of typical outgoing travel distance for each of the eight Mezuro regions. The measure therefore captures differences in between-region mobility rather than movements occurring within a Mezuro region.

Outgoing mobility rate was used as a supplementary measure of mobility intensity and as an alternative mobility indicator in sensitivity analyses. It was defined as the daily number of observed outgoing movements relative to the population size of each Mezuro region, averaged across the study period. As with average outgoing distance, this measure was available at the Mezuro-region level and therefore comprised eight independent regional observations. It may include multiple movements by the same individual within a day and does not capture within-region movement; it was consequently interpreted as a proxy for relative mobility intensity between regions rather than an individual-level travel rate.

#### 2.3.2 Health Outcome Indicators

Health outcomes were defined using complementary indicators of co-occurring disease burden. Co-occurring disease burden can be operationalised in different ways. Single-disease indicators may not adequately capture multimorbidity (Calderón-Larrañaga et al., 2019), while polypharmacy-based measures reflect accumulated medication use and are commonly used as proxies for overall health burden (Van Wilder et al., 2022; Gupta et al., 2025). Polypharmacy is typically defined as the concurrent use of five or more medications, although definitions vary (Masnoon et al., 2017; Pazan & Wehling, 2021). In contrast, syndemic frameworks emphasise interactions between co-occurring conditions within shared social and environmental contexts (Singer, 2009). These approaches may therefore capture different dimensions of population health.

In this study, cardiometabolic medication indicators were included as domain-specific complements to polypharmacy: while polypharmacy captures overall accumulated medication use, cardiometabolic medication prescriptions allow specific components of this broader medication burden to be examined separately. Cardiometabolic medication indicators were constructed from Anatomical Therapeutic Chemical (ATC) classification codes (World Health Organization, 2026). Medication records were classified according to ATC code prefixes. Lipid-lowering medication was defined as codes beginning with C10, glucose-lowering medication as codes beginning with A10B, and blood-pressure-lowering medication as codes beginning with C03, C07, C08, or C09.

Broader co-occurring disease burden was captured using polypharmacy and a syndemic-based measure. Polypharmacy was defined as the proportion of individuals using five or more unique medications (≥5 ATC-4 codes), with a stricter threshold of ten or more medications (≥10 ATC-4 codes) examined in sensitivity analyses.

The syndemic-based measure was constructed using predefined combinations of conditions identified in a related analysis of ELAN data as showing evidence of additive interaction in relation to healthcare use and costs (Dubbeldeman et al., 2026). The combinations are listed in Appendix B. These combinations were derived using the Relative Excess Risk due to Interaction (RERI), which quantifies whether the joint effect of two conditions exceeds the expected sum of their individual effects. Individuals were classified as having a syndemic pattern if they had at least one of these predefined interacting condition combinations. For each neighbourhood, the syndemic measure was then calculated as the proportion of registered ELAN patients with at least one such combination. These proportions are therefore based on individuals registered with general practices participating in ELAN, rather than the full underlying population. In 2019, ELAN-covered patients represented on average 27.2% of The Hague residents, with neighbourhood coverage ranging from 10.8%-49.1%.

#### 2.3.3 Contextual Covariates

Contextual covariates were grouped into conceptual domains representing key dimensions of urban environment and population structure. These domains included age composition, socioeconomic position, education, migration background, housing, lifestyle and social factors, access to facilities like supermarkets, physical environment, and social vulnerability.

This categorisation was shaped by social determinants of health and syndemic frameworks. Established frameworks commonly distinguish domains such as economic stability, education, neighbourhood and built environment, healthcare access, and social and community context (Office of Disease Prevention and Health Promotion, n.d.). These contextual domains have been widely studied in relation to disease risk and health inequalities, including in cardiovascular and cardiometabolic research (Powell-Wiley et al., 2022; Tamura et al., 2019).

Inclusion of migration background was motivated by evidence that health risks and mobility-related behaviours differ across population groups, with extensive evidence suggesting that these differences reflect social, cultural, structural, and life-course processes (Agyemang et al., 2024; Agyemang & Van den Born, 2022; Malmusi et al., 2010; Lebano et al., 2020; Loi et al., 2025). Housing, facilities access, and the physical environment were included because built-environment characteristics can shape health inequalities through pathways such as physical activity opportunities, access to services, environmental exposures, and material resources (Sims et al., 2020; Smith et al., 2017; Liu et al., 2023b).

Social vulnerability indicators were included to capture area-level disadvantage and susceptibility to adverse health outcomes not fully represented by income and education alone. These indicators reflected economic insecurity, debt-related vulnerability, and several other adverse social conditions. This approach is consistent with social vulnerability frameworks, which conceptualise vulnerability as a multidimensional construct arising from accumulated social, economic, household, and support-related disadvantages (Flanagan et al., 2011; Mah et al., 2023).

Multiple domain candidate variables were considered for further analysis (Table 1). The domains were used throughout the analysis to structure variable selection, assess contextual explanations of mobility, and define adjustment sets for the neighbourhood-level association analyses.

**Table 1.** Conceptual domains and examples of candidate contextual variables considered for variable selection.

| Domain | Candidate variables/examples |
| --- | --- |
| Age composition | Proportion aged 0-20, 21-40, 41-60, 61-80 |
| Socioeconomic position | Low income, insurance arrears |
| Education | Low, medium, high educational attainment |
| Migration background | Western/non-Western migration background, Statistics Netherlands-defined origin groups |
| Housing | Multi-family housing, owner-occupied housing |
| Lifestyle and social factors | Adherence to physical activity guidelines, social isolation |
| Facilities access | Distance to supermarket, facilities within 3 km |
| Physical environment | Public green space, PM<sub>10</sub> and NO<sub>2</sub> concentrations |
| Social vulnerability | Welfare benefit use, victim support clients |

### 2.4 Analytical Strategy

The analytical framework used in this study is illustrated in Figure 2. It summarises the process of variable selection, domain construction, contextual modelling of mobility, and residual-based association analysis.

**Figure 2.**
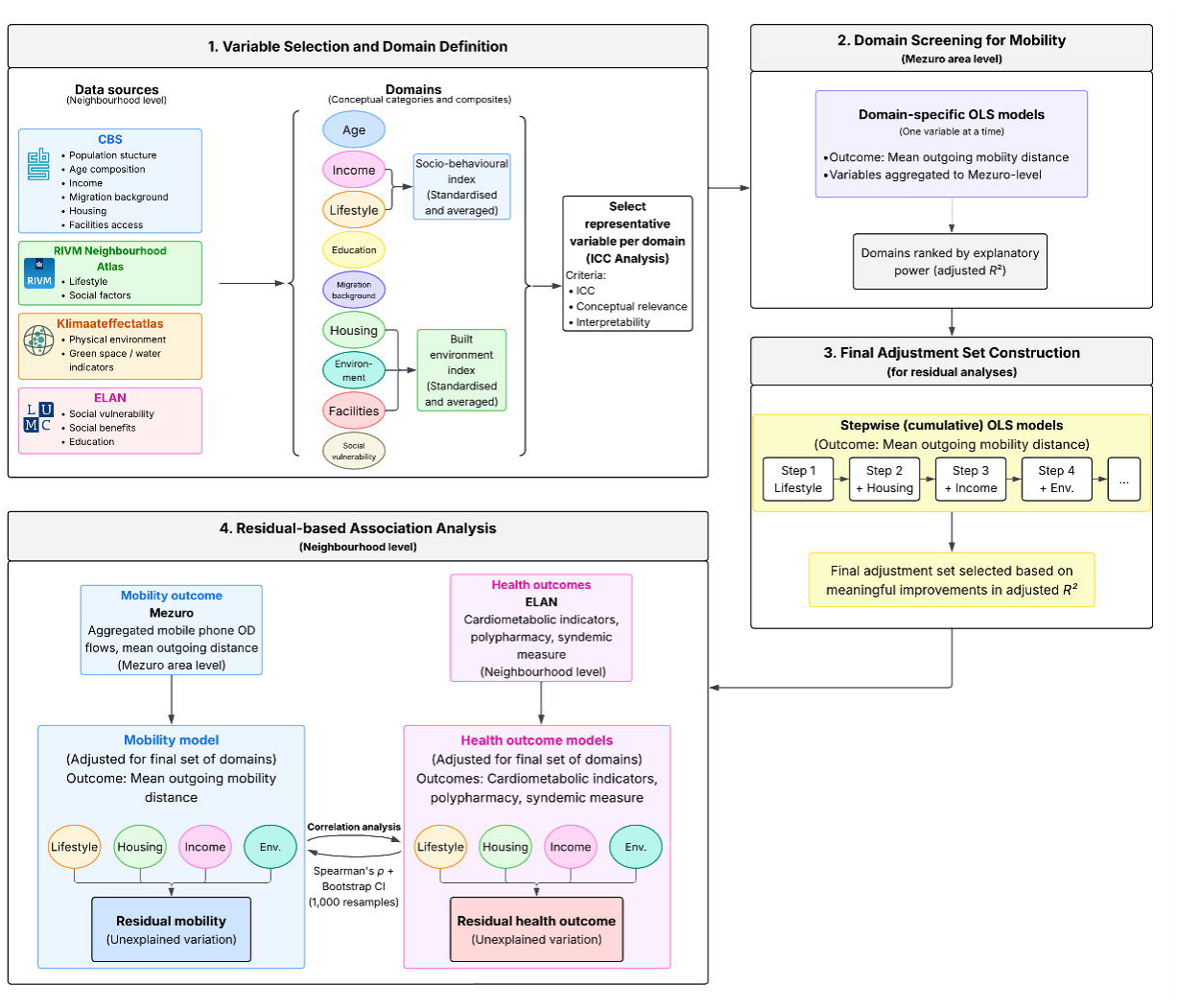
Analytical framework. Contextual variables were grouped into domains and screened using intra-class correlation (ICC) analysis to select representative covariates. Domain-specific and cumulative Ordinary Least Squares (OLS) models were used to explain regional mobility and define an adjustment set. Residuals from models of mobility and health outcome models were then correlated to assess whether mobility captures variation in health beyond shared contextual factors.

Contextual variables were grouped into conceptual domains and screened using intra-class correlation (ICC). Selected variables were subsequently examined in OLS models of mobility and used to construct a parsimonious contextual adjustment set. Residual mobility–health associations were then assessed at both Mezuro-region and neighbourhood levels, accounting for the cross-scale structure of the data.

#### 2.4.1 Variable Selection

To assess the degree of spatial clustering in candidate contextual covariates ICC-coefficients were calculated for all contextual variables. Formally, ICC was defined as the proportion of total variance attributable to differences between Mezuro regions:

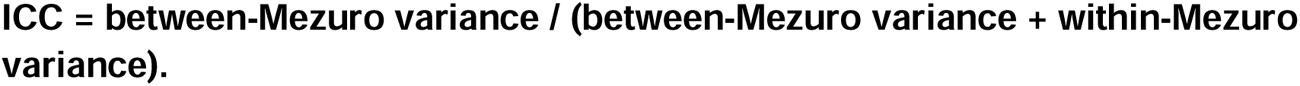

Higher ICC values indicate that a greater share of variation occurs between Mezuro regions and is therefore spatially structured at a scale more comparable to the regional mobility measure. Within each conceptual domain, ICC was considered alongside theoretical relevance, interpretability, and comparability with the mobility data. One representative variable per domain was selected, reducing dimensionality given the eight independent Mezuro mobility regions.

Intraclass correlation coefficients were calculated for all candidate contextual variables. Figure 3 presents the three highest-ranking variables within each domain, while a more extensive overview of up to ten variables per domain is provided in Appendix C (Table C1). Table C1 also identifies the variables selected for primary and sensitivity analyses.

#### 2.4.2 Contextual Correlates of Mobility

To examine which contextual domains were most strongly associated with regional mobility patterns, domain-specific OLS models were estimated at the Mezuro-region level. Given the limited number of Mezuro regions, models simultaneously including multiple contextual domains were not considered appropriate. Instead, each selected contextual variable was analysed separately, and adjusted R² was used descriptively to compare the explanatory contribution of the different domains to variation in outgoing mobility.

In addition, composite indices were constructed to represent broader contextual dimensions. Component variables were standardised to z-scores and averaged with equal weighting. Where necessary, variables were reverse-coded before averaging so that higher values consistently represented the same conceptual direction within each index. The socio-behavioural index combined indicators of socioeconomic disadvantage and lifestyle, whereas the built-environment index combined housing, physical-environment, and facilities-access indicators. These indices were analysed descriptively in the same manner as the individual domain variables to assess their association with regional differences in mobility.

#### 2.4.3 Adjustment Strategy for Neighbourhood-level Analyses

To define a minimal covariate set for neighbourhood-level analyses, contextual domains were added cumulatively based on their explanatory contribution to mobility. Domains were first ranked according to their adjusted *R*² in domain-specific models. Cumulative models were then evaluated by sequentially adding domains and assessing changes in adjusted *R*². Domains that provided meaningful improvements in variance explanation were retained in the final adjustment set.

#### 2.4.4 Association Analysis

Association between adjusted mobility and residual disease-burden indicators was assessed using complementary analyses at the Mezuro-region and neighbourhood levels. Adjusted mobility was defined as the residual from the OLS model in which mean outgoing mobility distance was modelled as a function of the selected contextual domains. Residual disease-burden indicators were defined as the residuals from separate OLS models in which each health outcome indicator was modelled using the same contextual domains.

First, a conservative region-level analysis was performed to ensure that the spatial resolution of the mobility data was fully respected. Adjusted mobility and disease-burden residuals were averaged within each Mezuro region, and Spearman’s rank correlation coefficient (ρ) was calculated across the eight Mezuro regions. Given the small number of regions, these correlations were interpreted descriptively.

Second, cross-scale associations were assessed using the neighbourhood-level residuals. Spearman’s coefficient was used to quantify the association between residual mobility and each residual disease-burden indicator. Although these analyses used neighbourhood-level residuals, the underlying mobility measure was observed for only eight Mezuro regions, and neighbourhoods within the same Mezuro region therefore did not represent independent mobility observations.

To account for this clustered structure when estimating uncertainty, 95% confidence intervals were obtained using 1,000 cluster-bootstrap resamples. Entire Mezuro regions, including all neighbourhoods belonging to each selected region, were resampled with replacement rather than resampling neighbourhoods independently. This preserved the dependence of neighbourhood observations within Mezuro regions and treated the eight Mezuro regions as the independent resampling units.

In addition, leave-one-region-out (LORO) analyses were performed to assess the influence of individual Mezuro regions. The cross-scale Spearman correlation was recalculated eight times, each time excluding one Mezuro region and all neighbourhoods belonging to it. The range of these correlations was used to evaluate whether the estimated association was strongly influenced by any single region.

Analyses were conducted for cardiometabolic medication indicators, polypharmacy, and the syndemic-based measure, allowing assessment of whether mobility–health associations differed across alternative conceptualisations of co-occurring disease burden.

#### 2.4.5 Sensitivity Analyses

Sensitivity analyses assessed robustness to alternative modelling choices. First, analyses were repeated using outgoing mobility rate instead of mean outgoing distance. Second, polypharmacy was redefined using a stricter threshold of ten or more medications (≥ 10 unique ATC4 codes in a year). Third, selected contextual variables were substituted with alternative indicators from the same domains. These substitutes were chosen because they represented the same broader conceptual domain, such as socioeconomic disadvantage, facilities access, or physical environment, while providing a different aspect of that domain. ICC values were also considered to ensure that substitute variables captured meaningful between-region variation. Together, these analyses evaluated whether mobility–health associations were stable across alternative mobility measures, multimorbidity definitions, contextual variable selections, and approaches to population composition.

## 3. Results

Table 2 presents descriptive statistics of mobility patterns and disease-burden indicators across the eight Mezuro regions. Clear regional variation was observed. Mean daily outgoing mobility distance ranged from 9.5 to 12.7 km, while daily outgoing mobility rates ranged from 71.2% to 94.0%. The two mobility measures seem to capture different dimensions of regional mobility: regions with longer average outgoing travel distances generally showed lower outgoing movement rates (Spearman’s ρ = −0.81).

**Table 2.** Descriptive statistics of mobility patterns and disease-burden indicators across eight Mezuro regions. Values are reported as mean (μ) and standard deviation (σ) calculated from neighbourhood-level observations within each region. The “Across-region” column reflects between-region variation (mean and standard deviation across Mezuro regions).

| Variable | Centrum | Escamp | Kijkduin | Oost | Scheveningen | West | Ypenburg | Zuid | Across-region mean (SD) |
| --- | --- | --- | --- | --- | --- | --- | --- | --- | --- |
| Population (N) | 33,535 | 92,040 | 43,850 | 48,240 | 66,500 | 111,305 | 48,690 | 93,600 | **67,220 (28,386)** |
| Outgoing distance (km), $\mu$ ($\sigma$) | 12.7 (17.5) | 9.5 (11.9) | 9.6 (10.7) | 12.6 (15.7) | 11.8 (15.7) | 11.7 (16.7) | 9.7 (10.8) | 10.5 (13.8) | **11.0 (1.36)** |
| Outgoing rate (%), $\mu$ ($\sigma$) | 71.2 (2.1) | 88.5 (3.1) | 75.4 (2.6) | 73.3 (2.1) | 74.2 (2.3) | 82.2 (2.0) | 94.0 (2.1) | 85.1 (2.1) | **80.5 (8.2)** |
| Cholesterol medication (%), $\mu$ ($\sigma$) | 7.6 (0.9) | 11.9 (1.5) | 14.7 (3.4) | 9.6 (2.7) | 9.5 (1.7) | 11.0 (2.8) | 9.1 (0.1) | 10.6 (2.8) | **10.6 (2.6)** |
| Diabetes medication (%), $\mu$ ($\sigma$) | 2.9 (0.9) | 5.9 (1.0) | 4.8 (1.4) | 3.1 (1.0) | 2.9 (1.0) | 4.7 (1.8) | 3.6 (0.3) | 6.3 (2.3) | **4.5 (1.4)** |
| Blood pressure medication (%), $\mu$ ($\sigma$) | 11.5 (0.9) | 16.0 (1.5) | 22.3 (4.5) | 17.2 (4.7) | 15.5 (2.6) | 16.6 (5.1) | 12.6 (0.0) | 13.5 (2.7) | **15.5 (3.7)** |
| Polypharmacy $\geq 5$ meds (%), $\mu$ ($\sigma$) | 16.2 (1.7) | 25.4 (3.0) | 26.4 (5.5) | 21.2 (5.1) | 18.5 (3.1) | 22.7 (5.1) | 17.8 (1.2) | 23.2 (6.2) | **21.6 (4.2)** |
| Syndemic measure (%), $\mu$ ($\sigma$) | 7.3 (5.1) | 17.3 (7.9) | 15.3 (7.7) | 13.5 (8.5) | 14.7 (21.2) | 12.9 (6.7) | 7.4 (6.5) | 7.5 (5.6) | **10.2 (4.6)** |

Medication-based disease indicators also varied across regions. Cholesterol medication use ranged from 7.6% to 14.7%, diabetes medication use from 2.9% to 6.3%, and blood pressure medication use from 11.5% to 22.3%. Multimorbidity measures showed comparable variation, with polypharmacy (≥5 medications) ranging from 16.2% to 26.4%, and the syndemic measure from 7.3% to 17.3%. These descriptive results indicate meaningful regional differences in both mobility and disease burden.

The regions with the highest and lowest values were not identical across the disease-burden indicators, suggesting that the spatial patterning of disease burden depends on the outcome measure used. For example, Mezuro region Kijkduin showed highest levels of cholesterol medication use, blood pressure medication use, whereas Zuid and Escamp showed highest diabetes medication use and syndemic burden. This indicates that different disease-burden indicators capture partly distinct dimensions of population health.

### 3.1 Variable Selection

**Figure 3.**
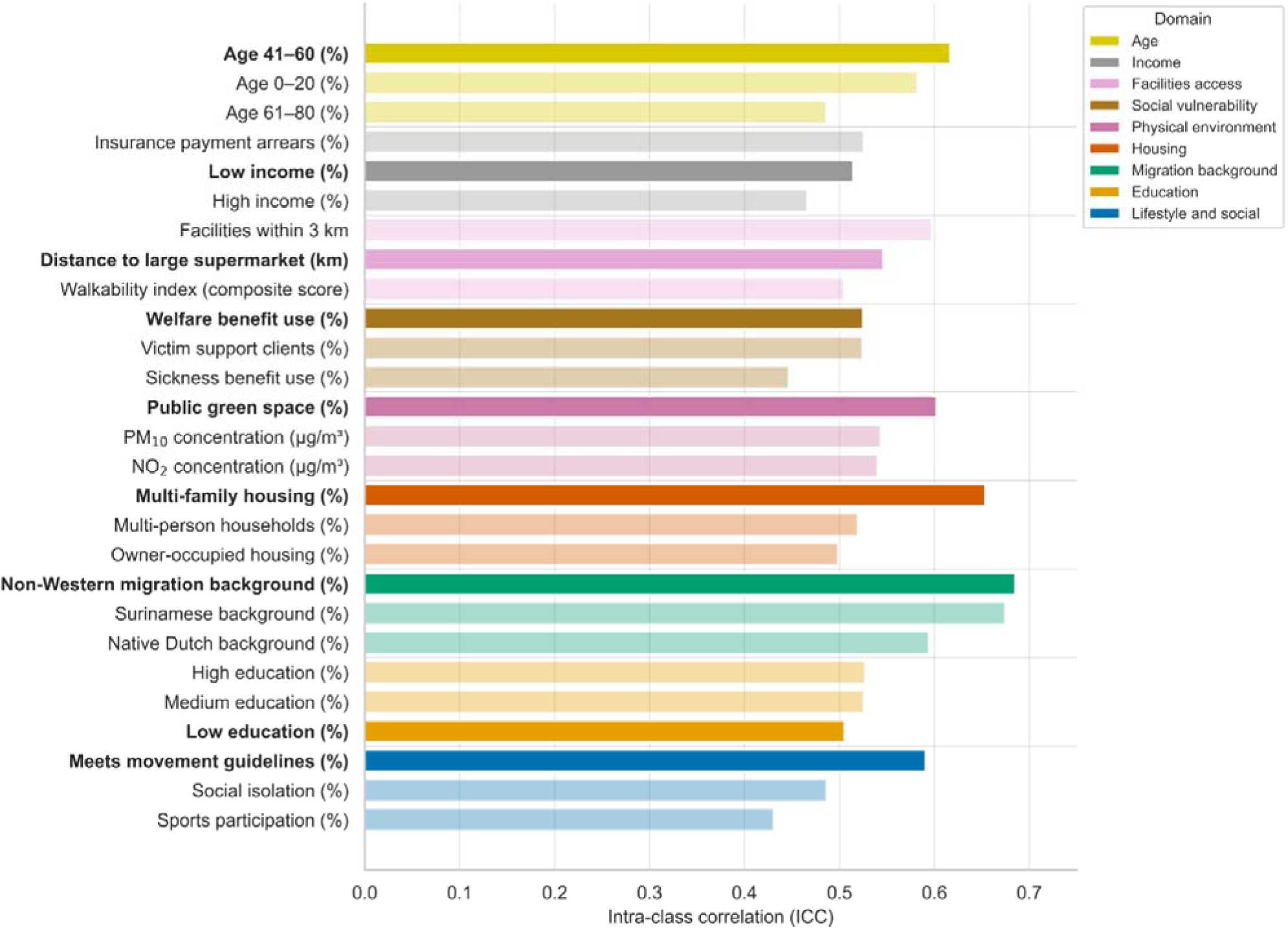
Top three contextual variables per domain ranked by intra-class correlation (ICC). Higher ICC values indicate stronger spatial clustering at the Mezuro-region level. Variables selected for subsequent analyses are highlighted in bold.

Figure 3 presents the three highest-ranking contextual variables within each domain based on ICC. A more extensive overview of the ICC rankings, including up to ten candidate variables per domain, is provided in Appendix C (Table C1). Substantial between-region variation was observed across multiple domains. Variables related to migration background showed strongest difference, with non-Western migration background having the highest ICC (0.68), followed by Surinamese background (0.67). Housing characteristics were also strongly spatially structured, particularly multi-family housing (ICC = 0.65).

Demographic, lifestyle, educational, environmental, facilities-access, social vulnerability, and income variables also showed meaningful between-region variation. For example, the proportion of individuals aged 41-60 (0.62), adherence to physical activity guidelines (0.60), public green space (0.60), and welfare benefit use (0.52) all demonstrated moderate to strong between-area variance.

Selected variables are highlighted in bold in Figure 3. As explained before, selection was not based solely on the highest ICC within each domain. For example, low educational attainment was selected instead of high education to capture disadvantage, distance to supermarkets was preferred over facility density as it provided a more direct measure of access and was available at a more appropriate spatial scale, and low income was selected over insurance arrears as a broader indicator of socioeconomic position.

### 3.2 Contextual Correlates of Regional Mobility Patterns

Domain-specific OLS models showed substantial variation in how well contextual domains explained outgoing mobility at the Mezuro-region level. Lifestyle showed the strongest association, with adherence to physical activity guidelines explaining the largest variance share (adjusted R² = 0.742). Education also showed substantial explanatory power (adjusted R² = 0.493), while multi-family housing and public green space explained moderate variance proportions (adjusted R² = 0.202 and 0.195, respectively).

Composite indices showed modest explanatory power. The socio-behavioural index explained 20.3% of variance in mobility, while the built environment index explained 12.4%. In contrast, several variables with strong between-area variance showed limited explanatory relevance for mobility. For example, non-Western migration background explained little variation in outgoing mobility (adjusted R² = 0.034), indicating that high ICC, meaning strong clustering of a variable between Mezuro regions, does not necessarily translate into explanatory power for mobility.

In an exploratory diagnostic analysis, Western migration background was strongly associated with outgoing mobility at the Mezuro-region level (ρ = 0.72; adjusted R² = 0.893). This indicates that Mezuro regions with a higher proportion of residents with a Western migration background had longer average outgoing mobility distances. Because both variables were aggregated at the Mezuro-region level and only eight regions were available, this result should be interpreted descriptively as an area-level compositional association, not as evidence of individual travel behaviour. Additional diagnostic information, including region-level values and scatterplots, is provided in Appendix E.

### 3.3 Neighbourhood-level Associations Between Mobility and Health Outcomes

Contextual domains were sequentially added based on their descriptive contribution to outgoing mobility and their conceptual relevance. Lifestyle showed the strongest association with outgoing mobility, while housing, environment, and income further improved model fit. These domains were therefore selected as the primary adjustment set. The cumulative models in Table 3 were fitted at the neighbourhood level to construct the context-adjusted mobility measure used in the subsequent cross-scale analysis. Because observed mobility varied only across eight Mezuro regions and was shared by neighbourhoods within each region, these model-fit statistics were used descriptively rather than for conventional neighbourhood-level statistical inference. Table 3 shows only retained steps; the full stepwise inclusion process is provided in Appendix D.

**Table 3.** Descriptive cumulative model fit for the cross-scale neighbourhood-level adjustment model of outgoing mobility. Because mobility was observed for eight Mezuro regions and assigned to their constituent neighbourhoods, adjusted R² values are presented as model-fit summaries rather than as evidence based on independent neighbourhood-level mobility observations.

| Model | Included domains | Adjusted $R^2$ |
| --- | --- | --- |
| 1 | Lifestyle | 0.388 |
| 2 | + Housing | 0.612 |
| 3 | + Environment | 0.659 |
| 4 | + Income | 0.732 |

After adjustment for lifestyle, housing, environment, and income, associations between residual mobility and residual disease-burden indicators were generally weak at the neighbourhood level (Table 4). Because mobility was independently observed for only eight Mezuro regions, uncertainty was estimated by resampling complete Mezuro regions rather than individual neighbourhoods. Neighbourhood-level Spearman correlations were close to zero for diabetes medication use (ρ = - 0.06), cholesterol medication use (ρ = -0.11), blood pressure medication use (ρ = - 0.04), polypharmacy (ρ = 0.01), and the syndemic measure (ρ = 0.10). Cluster-bootstrap 95% confidence intervals included zero for all outcomes and were relatively wide, reflecting the limited number of independent mobility regions.

**Table 4.** Associations between mobility and health outcomes after adjustment for contextual domains (lifestyle, housing, environment, and income). LORO ranges show the variation in the neighbourhood-level Spearman correlation when each Mezuro region was omitted in turn.

| Outcome | Adjusted R<sup>2</sup> | Mezuro-level $\rho$ | Neighbourhood $\rho$ [Cluster-bootstrap CI] | LORO neighbourhood $\rho$ range |
| --- | --- | --- | --- | --- |
| Diabetes medication | 0.796 | -0.26 | -0.06 [-0.45, 0.23] | -0.20 to 0.08 |
| Cholesterol medication | 0.338 | -0.45 | -0.11 [-0.33, 0.13] | -0.21 to -0.04 |
| Blood pressure medication | 0.210 | -0.38 | -0.04 [-0.30, 0.23] | -0.12 to 0.06 |
| Polypharmacy $\geq 5$ | 0.497 | -0.36 | 0.01 [-0.38, 0.36] | -0.15 to 0.10 |
| Syndemic measure | 0.065 | -0.48 | 0.10 [-0.18, 0.40] | -0.07 to 0.17 |

Mezuro-region level correlations were larger in magnitude, particularly for cholesterol medication use (ρ = -0.45), blood pressure medication use (ρ = -0.38), and the syndemic measure (ρ = -0.48). Taken together, these results suggest that the mobility-health relationship at the neighbourhood level was largely shared with the measured contextual conditions. Lifestyle, housing, physical environment, and income accounted for substantial variation in outgoing mobility in the descriptive adjustment model, while little association remained between the residual components of mobility and disease burden after these same domains were accounted for. Mobility therefore provided limited additional health-related information beyond the measured contextual structure at this spatial scale.

### 3.4 Sensitivity Analyses

Sensitivity analyses generally supported the primary findings. Replacing mean outgoing distance with outgoing mobility rate yielded similar results, with weak cross-scale neighbourhood associations and cluster-bootstrap confidence intervals including zero. Using a stricter polypharmacy definition (≥10 medications) also did not materially change the interpretation.

**Table 5.**
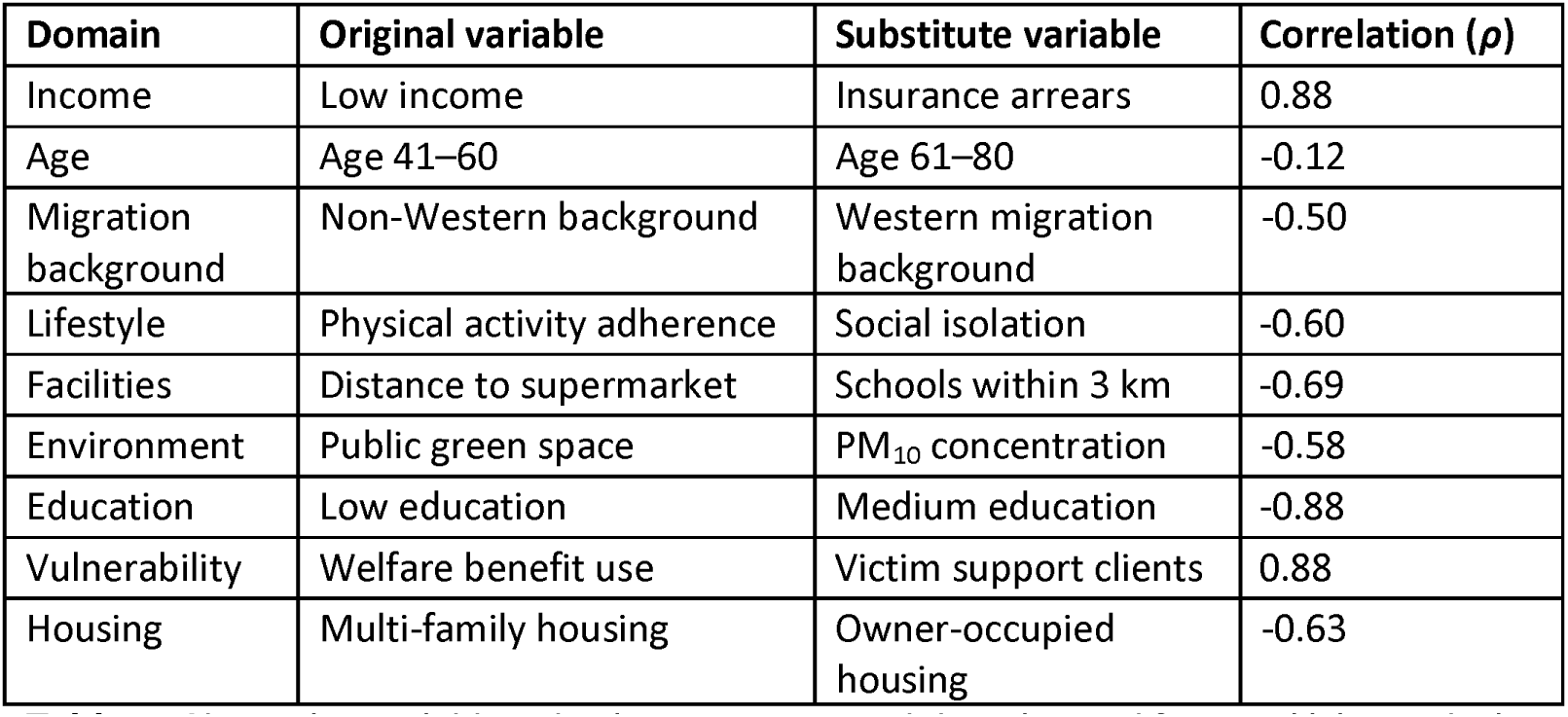
Alternative variable selection per contextual domain used for sensitivity analysis. Correlations indicate similarity between original and substitute variables.

| Domain | Original variable | Substitute variable | Correlation ($\rho$) |
| --- | --- | --- | --- |
| Income | Low income | Insurance arrears | 0.88 |
| Age | Age 41–60 | Age 61–80 | -0.12 |
| Migration background | Non-Western background | Western migration background | -0.50 |
| Lifestyle | Physical activity adherence | Social isolation | -0.60 |
| Facilities | Distance to supermarket | Schools within 3 km | -0.69 |
| Environment | Public green space | PM<sub>10</sub> concentration | -0.58 |
| Education | Low education | Medium education | -0.88 |
| Vulnerability | Welfare benefit use | Victim support clients | 0.88 |
| Housing | Multi-family housing | Owner-occupied housing | -0.63 |

Robustness to contextual variable selection was assessed by substituting alternative variables within each domain (Table 5). Several substitutes were strongly correlated with the original variables, although some correlations were negative because domain variables represented opposite directions, such as public green space versus air pollution or non-Western versus Western migration background.

Using alternative variables changed the ranking of domains modestly. Migration background, operationalised as Western migration background, showed a stronger descriptive cross-scale association with mobility at the neighbourhood level (adjusted R² = 0.498). Within the alternative-variable specification on the neighbourhood-level, inclusion of Western migration background increased the descriptive adjusted R² of the neighbourhood-level mobility model from 0.346 to 0.514. As in the primary analysis, these model-fit statistics were not interpreted using conventional neighbourhood-level inference because mobility was independently observed for only eight Mezuro regions. Despite these differences in variable specification, cross-scale mobility–health associations remained small across outcomes when using the alternative adjustment set (Table 6).

At the Mezuro-region level, descriptive Spearman correlations remained negative across all outcomes, ranging from ρ = −0.19 for polypharmacy ≥10 medications to ρ = −0.36 for diabetes and cholesterol medication use. In contrast, cross-scale neighbourhood correlations after alternative contextual adjustment were small, ranging from ρ = −0.11 to 0.08. Cluster-bootstrap 95% confidence intervals included zero for all outcomes and were relatively wide.

LORO-analyses similarly showed that the cross-scale associations were not strongly dependent on any single Mezuro region. For diabetes and cholesterol medication use, for example, correlations remained between −0.20 and 0.06 when individual regions were excluded. Across the remaining outcomes LORO-estimates were also small, although the direction of the association varied for several outcomes. Together, these analyses indicate that the finding of weak cross-scale mobility– health associations was robust to alternative contextual variable selection and was not strongly driven by any single Mezuro region.

**Table 6.**
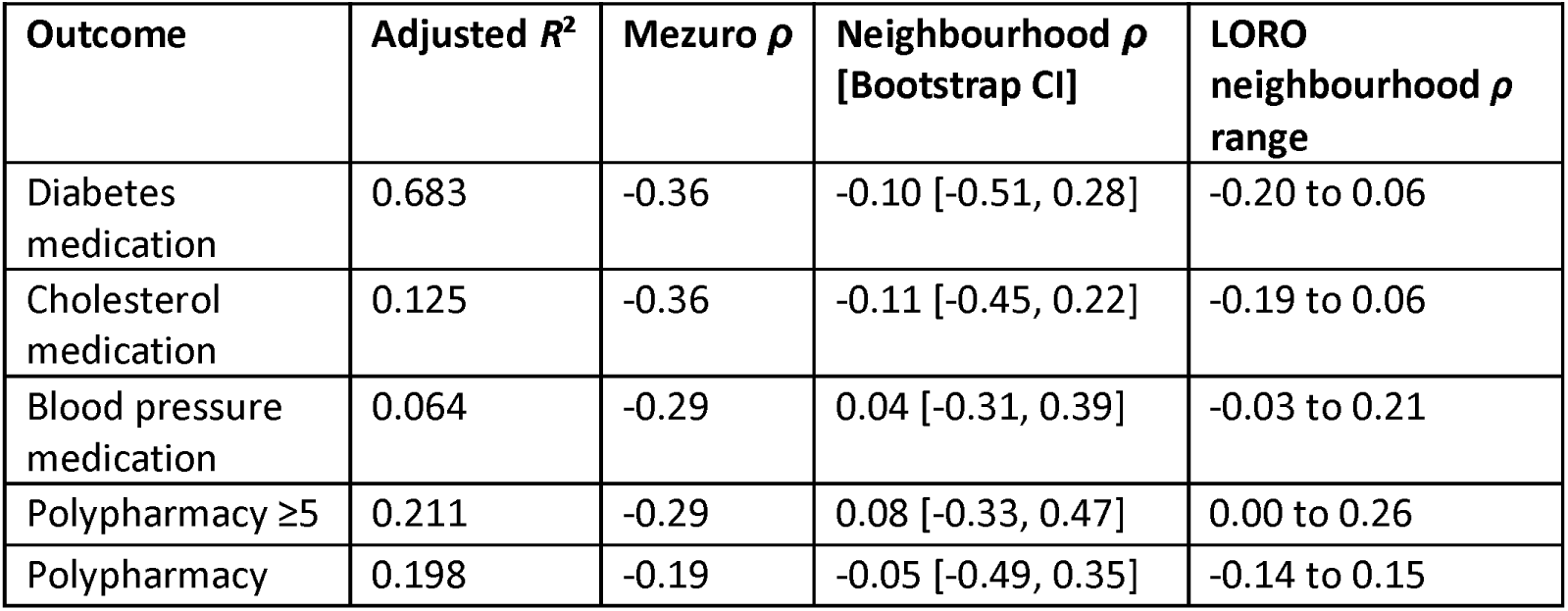
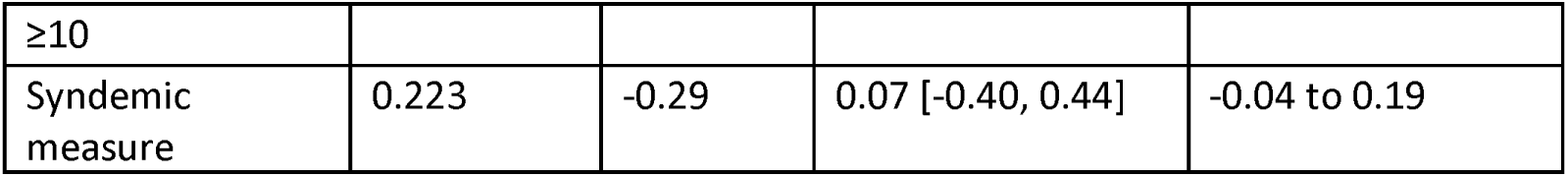
Sensitivity analysis of the association between mobility and health outcomes using alternative contextual variables. Both mobility and the outcomes were corrected for migration background, lifestyle, housing, environment, and income. Associations between mobility and health outcomes remain weak, with cluster-bootstrap confidence intervals including zero.

Because Western migration background was strongly associated with outgoing mobility, an additional sensitivity analysis was conducted at the Mezuro-region level. Detailed results are provided in Appendix E. Because both outgoing mobility and Western migration background were defined at the Mezuro-region level, these estimates primarily reflect between-region variation. They were therefore treated as diagnostic sensitivity results rather than primary neighbourhood-level findings.

## 4. Discussion

This study examined how aggregated urban mobility relates to population health and contextual conditions across different spatial supports. Outgoing mobility was strongly patterned by urban context, while residual mobility–health associations were weak in the cross-scale neighbourhood analyses after accounting for shared contextual conditions. This pattern suggests that aggregated mobility primarily reflected contextual characteristics that were themselves related to population health, rather than providing substantial additional health-related information as a standalone indicator. In other words, areas with different mobility patterns also differed systematically in the social, behavioural, and environmental conditions considered in this study; after these differences were accounted for, little neighbourhood-level mobility-health association remained.

### 4.1 Spatial Scale and the Interpretation of Mobility-Health Relationships

The findings highlight the importance of spatial scale and geographic support when interpreting mobility–health relationships. At the Mezuro-region level, longer outgoing mobility distances were moderately associated with lower disease burden, whereas these relationships were substantially weaker after contextual adjustment in the cross-scale analyses. Mobility was measured for eight relatively extended Mezuro regions, while contextual characteristics and health outcomes were available for finer administrative neighbourhoods. The contrast therefore likely reflects both contextual adjustment and the limited ability of a regional mobility measure to represent health variation between neighbourhoods within the same region.

This distinction also relates to broader geographical critiques of relying exclusively on residential neighbourhoods to understand population exposures. Activity-space and spatial-polygamy approaches emphasise that people interact with multiple environments through daily travel and are therefore not exposed solely to conditions within their residential neighbourhood (Matthews & Yang, 2013; Chaix, 2018). Mobile-phone-derived mobility data offer an opportunity to represent some of these connections between places. However, the present findings demonstrate that aggregated inter-regional mobility is not equivalent to an individual activity space. Mean outgoing distance characterises the spatial reach of movements originating from a region, but does not identify individual destinations, trip purposes, transport modes, or the environments encountered during travel. Regional mobility may therefore capture urban connectivity while poorly representing finer-scale exposure processes relevant to health.

This does not imply that aggregated mobility data are inherently too coarse for population-health research. Their usefulness depends on whether the scale and meaning of the mobility measure correspond to the process studied. Aggregated mobile-phone mobility has been particularly useful for infectious-disease dynamics, where flows between larger areas are directly relevant (Turnnidge et al., 2025; Grantz et al., 2020; Kostandova et al., 2024). Chronic disease burden, by contrast, reflects processes operating across multiple spatial and temporal scales.

Taken together, these findings show that mobility-health associations depend on contextual confounding and on the geographical units through which mobility and health are represented. Spatial resolution should therefore be treated as part of the substantive interpretation of mobility data rather than solely as a technical limitation.

### 4.2 Population composition and the interpretation of aggregated mobility

Western migration background was strongly associated with outgoing mobility distance at the Mezuro-region level, accounting for 89.3% of its variance. This area-level association was robust to population weighting and was not driven by a single region, but does not imply that individuals with a Western migration background travel farther. Rather, it illustrates how aggregated mobility may simultaneously reflect population composition, characteristics of place, and spatial connectivity, all of which are geographically structured within the city.

The association was particularly apparent for outgoing mobility distance and was not accompanied by a comparable pattern in mobility volume. This suggests that the spatial reach of movements may be especially sensitive to population composition and the geographical organisation of the city. Previous research has reported differences in mobility behaviour between population groups in the Netherlands (Durand et al., 2023; Delbosc & Shafi, 2023; Faber et al., 2025), but these individual-level findings cannot be used to explain the regional association observed here directly. Instead, the present results indicate that population composition should be considered when interpreting geographical variation in aggregated mobility.

The migration-background analysis was therefore treated as exploratory and addressed a different question from the primary adjustment strategy. Whereas the primary analysis assessed whether mobility retained health-relevant variation after accounting for several contextual domains simultaneously, adjustment for Western migration background isolated the influence of one particularly strong area-level compositional variable. Its importance lies in demonstrating how strongly the interpretation of an aggregated mobility indicator can depend on the composition of the populations represented by the spatial units, rather than identifying migration background as an explanatory mechanism. Together with the spatial-scale findings, this supports interpreting aggregated mobility as a hybrid indicator of regional context, population composition, and connectivity rather than as a direct measure of individual mobility behaviour.

### 4.3 Strengths and Limitations

This study has several strengths. First, it combines large-scale mobility data with very detailed contextual and health information within a multi-scale analytical framework. Importantly, the analysis explicitly accounted for the different spatial supports at which mobility, contextual characteristics, and health outcomes were observed. Mezuro-region level analyses were used to characterise regional mobility patterns, while neighbourhood-level analyses were used to assess associations between residual mobility and residual chronic disease-burden indicators. Cluster-bootstrap confidence intervals and leave-one-region-out analyses were additionally used to reflect the clustered spatial structure of the data.

Second, the residual-based modelling approach enabled a more nuanced interpretation of mobility than a direct association analysis. Rather than treating mobility as an isolated exposure, the analysis tested whether mobility retained an association with health outcomes after accounting for selected contextual domains. This multidimensional approach is particularly relevant because aggregated mobility may simultaneously reflect characteristics of the population, characteristics of place, and connectivity to the wider urban system. This is compatible with a syndemic-informed perspective because it considers shared social and environmental conditions that may shape disease burden, while also showing that residual mobility was not independently associated with the syndemic-based disease burden measure.

Third, this study used complementary indicators of co-occurring disease burden. In addition to individual medication indicators for glucose-, cholesterol-, and blood pressure control, both polypharmacy thresholds and a syndemic-based measure were included. This allows for the assessment of mobility–health associations across different dimensions of disease burden, including both the accumulation and interaction of conditions. Such a multidimensional approach strengthens the robustness of conclusions and provides a broader understanding of population health patterns.

Fourth, extensive sensitivity analyses were conducted to evaluate robustness of findings under different modelling assumptions. These included use of alternative mobility measures, different definitions of multimorbidity, and substitution of contextual variables within domains. Importantly, we explicitly examined the role of migration background as a dominant population-compositional factor influencing mobility. By demonstrating how results change when adjusting for Western migration background, and by showing that this association primarily concerned outgoing mobility distance rather than mobility volume, the analysis provides valuable insight into how aggregated mobility indicators can reflect population composition, spatial reach, and contextual structure simultaneously.

Several limitations should be considered. First, mobility was measured using aggregated origin-destination flows at the Mezuro-region level and summarised over the study period. Most importantly, mobility was independently observed for only eight Mezuro regions, which substantially limited statistical power and the complexity of region-level models that could be estimated reliably. Region-level associations were therefore interpreted descriptively, and the neighbourhood-level analyses should be understood as cross-scale analyses rather than as analyses based on 44 independent mobility observations. Although cluster-bootstrap and leave-one-region-out analyses better reflected this dependence structure, uncertainty estimates based on only eight clusters are inherently imprecise.

Although neighbourhood-level analyses increased the number of health observations, all neighbourhoods within the same Mezuro region were assigned the same mobility value. This spatial scale mismatch limited our ability to capture within-region heterogeneity and may have introduced exposure misclassification at neighbourhood level. The linkage between provider-defined mobility regions and administrative neighbourhoods also involved a change of spatial support, and neighbourhoods intersecting more than one Mezuro region were assigned according to greatest geographic overlap. This necessarily simplifies spatial relationships near regional boundaries. At the same time, this mismatch illustrates a broader challenge when mobility datasets produced using operational spatial units are combined with routinely available administrative statistics.

In addition, the daily aggregated provided data reduced temporal variation. The analysis therefore could not distinguish individual movement patterns, transport mode, trip purpose, active transport, commuting versus leisure travel, or activity spaces. These forms of aggregation may have weakened health-relevant variation and limited the ability to detect associations at finer spatial, temporal, or behavioural scales. Average outgoing distance should therefore be interpreted as an area-level measure of spatial reach between Mezuro regions rather than as a direct representation of individual mobility behaviour.

Second, mobility data were derived from the VodafoneZiggo network and therefore reflect mobility patterns of users connected to this provider network rather than all mobile-phone users. During the study period, the Vodafone network included Vodafone and a limited number of associated lower-cost or virtual providers, whereas several other budget or virtual providers operated on different networks. This provider-specific market coverage may have introduced selection bias if subscription choices differed by socioeconomic position, age, migration background, digital access, or neighbourhood.

Third, sensitivity analyses involving Western migration background highlight an important methodological challenge. Because this variable explains a very large proportion of variance in mobility, adjusting for it substantially alters the interpretation of the residual mobility measure. While this analysis provides valuable insight into the compositional nature of mobility, resulting associations should be interpreted as conditional and exploratory rather than confirmatory. Rather than contradicting the primary analysis, these findings add nuance by showing that aggregated mobility reflects both contextual conditions and regional population composition. All associations were estimated at area level and should not be extrapolated to individual behaviour or individual disease risk. This is particularly relevant for this migration-background analysis, where regional population composition was associated with mobility patterns but does not imply corresponding individual-level mobility differences.

## 5. Conclusion

This study examined how aggregated urban mobility relates to population health across different spatial scales and whether mobility retains population health-relevant information beyond contextual conditions. Mobility patterns varied substantially across Mezuro regions and were strongly associated with contextual and population-composition factors. Moderate mobility-health associations were observed at the regional level, but these relationships were substantially weaker in the cross-scale neighbourhood analyses after adjustment for key contextual domains. Residual mobility was not consistently associated with indicators of disease burden, including cardiometabolic medication prescriptions, polypharmacy, or syndemic-based disease burden.

These findings suggest that aggregated mobility should not be interpreted as a straightforward standalone indicator of population health. Rather, mobility appears to reflect broader social, environmental, behavioural, and demographic conditions that are themselves related to disease burden. The contrast between regional and neighbourhood-level findings further demonstrates that the interpretation of mobility-health relationships depends on the spatial support at which mobility and health are measured. The strong association with Western migration background additionally illustrates that aggregated mobility captures characteristics of the populations represented by the respective regions, next to regional spatial connectivity.

Overall, this study highlights both the promise and complexity of using mobility data in urban health and geographical research. Aggregated mobility can provide valuable insight into spatial connectivity and the multidimensional structure of urban areas, but its interpretation depends strongly on scale, aggregation, and population composition. When mobility data are combined with administrative health and contextual statistics, differences between the geographical units represented by these data should therefore form part of the substantive interpretation rather than being treated solely as a technical limitation. More granular mobility data, including transport mode, trip purpose, and individual activity spaces, could help clarify the mechanisms linking movement patterns to health. In this study, aggregated mobility was most informative as a spatially and contextually embedded indicator of urban connectivity and population composition, with limited additional association with neighbourhood disease burden after accounting for measured contextual conditions.

## Supporting information

Full Supplementary Material

## Declaration of generative AI and AI-assisted technologies in the manuscript preparation process

During the preparation of this work, the authors used ChatGPT to support language refinement. The authors reviewed and edited the content as needed and take full responsibility for the content of the submitted manuscript.

## Data availability statement

The area-level health data used in this study were obtained from the ELAN GGDH dashboard (https://elandcc.shinyapps.io/ELANDashboard/). The mobility data were obtained from Mezuro and are not publicly available because they are subject to data-use agreements. Publicly available contextual and environmental data were obtained from the RIVM Buurtatlas, the Klimaateffectatlas, and Statistics Netherlands’ 2019 neighbourhood data. Aggregated analysis outputs and code may be made available by the corresponding author upon reasonable request, where permitted by applicable data-use agreements.

## CRediT authorship contribution statement

**Hielke Muizelaar:** Conceptualisation, Investigation, Formal analysis, Visualization, Data curation, Writing – original draft, Writing – review & editing. **Marcel Haas**: Conceptualisation, Investigation, Supervision, Writing – review & editing. **Rimke Vos:** Conceptualisation, Investigation, Supervision, Writing – review & editing. **Ilonca Vaartjes**: Supervision, Writing – review & editing. **Ester de Jonge**: Supervision, Writing – review & editing. **Lampros Stergioulas**: Supervision, Writing – review & editing. **Jessica Kiefte-de Jong**: Conceptualisation, Investigation, Supervision, Project administration, Resources, Writing – review & editing. **Marco Spruit**: Conceptualisation, Investigation, Supervision, Project administration, Writing – review & editing.

## Acknowledgments

We thank the ECOTIP management team for their helpful comments and suggestions.

