## Supplementary material for "Urban Mobility and Population Health in The Hague: A Multi-Scale Analysis": Full Supplementary Material

### Appendix A: Neighbourhood assignment to Mezuro mobility regions

| No. | Neighbourhood | Assigned Mezuro mobility region | Overlap (%) |
| --- | --- | --- | --- |
| 1 | Archipelbuurt | Scheveningen | 87.5 |
| 2 | Belgisch Park | Scheveningen | 89.6 |
| 3 | Benoordenhout | Oost | 99.9 |
| 4 | Bezuidenhout | Oost | 98.9 |
| 5 | Binckhorst | Zuid | 100 |
| 6 | Bohemen en Meer en Bos | Kijkduin | 61.9 |
| 7 | Bomen- en Bloemenbuurt | West | 100 |
| 8 | Bouwlust | Escamp | 99.7 |
| 9 | Centrum | Centrum | 100 |
| 10 | Duindorp | Scheveningen | 100 |
| 11 | Duinoord | West | 93.1 |
| 12 | Forepark | Ypenburg | 100 |
| 13 | Geuzen- en Statenkwartier | Scheveningen | 94.3 |
| 14 | Groente- en Fruitmarkt | Zuid | 98.2 |
| 15 | Haagse Bos | Oost | 99.5 |
| 16 | Hoornwijk | Ypenburg | 99.7 |
| 17 | Kijkduin en Ockenburgh | Kijkduin | 99.8 |
| 18 | Kraayenstein en Vroondaal | Kijkduin | 100 |
| 19 | Laakkwartier en Spoorwijk | Zuid | 100 |
| 20 | Leidschenveen | Ypenburg | 100 |
| 21 | Leyenburg | West | 70.5 |
| 22 | Loosduinen | Kijkduin | 72.4 |
| 23 | Mariahoeve en Marlot | Oost | 100 |
| 24 | Moerwijk | Zuid | 99.5 |
| 25 | Morgenstond | Escamp | 88.2 |
| 26 | Oostduinen | Oost | 73.2 |
| 27 | Regentessekwartier | West | 72.6 |
| 28 | Rustenburg en Oostbroek | West | 100 |
| 29 | Scheveningen | Scheveningen | 100 |
| 30 | Schildersbuurt | Zuid | 80.8 |
| 31 | Stationsbuurt | Zuid | 87 |
| 32 | Transvaalkwartier | West | 96.9 |
| 33 | Valkenboskwartier | West | 100 |
| 34 | Van Stolkpark en Scheveningse Bosjes | Scheveningen | 95.2 |
| 35 | Vogelwijk | West | 63 |
| 36 | Vruchtenbuurt | West | 96.4 |
| 37 | Waldeck | Kijkduin | 98.3 |
| 38 | Wateringse Veld | Escamp | 99.9 |
| 39 | Westbroekpark en Duttendel | Oost | 72.8 |
| 40 | Willemspark | Centrum | 87.9 |
| 41 | Ypenburg | Ypenburg | 100 |
| 42 | Zeeheldenkwartier | Centrum | 80.7 |
| 43 | Zorgvliet | West | 99.9 |
| 44 | Zuiderpark | Zuid | 93.8 |

**Table A.1.** Administrative neighbourhoods shown in Figure 1C and their assigned Mezuro mobility region. Neighbourhoods were assigned to the Mezuro region with which they had the greatest geographic overlap (excluding the North Sea).

### Appendix B: Condition combinations used to construct the syndemic-based measure

This appendix lists the predefined condition combinations used to construct the syndemic-based measure. Combinations were derived from prior submitted, unpublished work by Dubbeldeman et al., in which additive interaction between health-condition dyads was assessed using the Relative Excess Risk due to Interaction (RERI). In that work, combinations showing statistically significant positive additive interaction were identified in sex- and age-stratified analyses. Individuals in the present study were classified as having a syndemic pattern if they had at least one of the condition combinations listed below in Table B.1.

| Sex | Age group | Health condition 1 | Health condition 2 | RERI | p-value |
| --- | --- | --- | --- | --- | --- |
| Female | 18–34 | Abdominal complaints/disorders | Skin disorders | 0.60 | 0.005 |
| Female | 18–34 | Mood and anxiety disorders | Abdominal complaints/disorders | 0.85 | <0.001 |
| Male | 18–34 | Mood and anxiety disorders | Respiratory infections | 3.18 | 0.002 |
| Female | 35–49 | Abdominal complaints/ disorders | Headache complaints | 0.90 | <0.001 |
| Female | 35–49 | Abdominal complaints/ disorders | Neck and back complaints | 0.91 | 0.002 |
| Female | 35–49 | Mood and anxiety disorders | Headache complaints | 0.78 | <0.001 |
| Female | 35–49 | Mood and anxiety disorders | Neck and back complaints | 1.01 | <0.001 |
| Female | 50–64 | Abdominal complaints/ disorders | Hypertension | 1.37 | 0.001 |
| Female | 50–64 | Hypertension | Cardiovascular diseases | 3.06 | <0.001 |
| Female | 50–64 | Hypertension | Mood and anxiety disorders | 1.08 | 0.005 |
| Female | 50–64 | Neck and back complaints | Mood and anxiety disorders | 1.51 | 0.003 |
| Male | 50–64 | Hypertension | Mood and anxiety disorders | 1.08 | 0.005 |
| Male | 50–64 | Hypertension | Neck and back complaints | 1.49 | 0.004 |
| Female | 65+ | Cardiovascular diseases | Abdominal complaints/disorders | 0.93 | <0.001 |
| Female | 65+ | Cardiovascular diseases | Asthma/ COPD | 0.83 | 0.001 |
| Female | 65+ | Cardiovascular diseases | Cancer | 0.59 | 0.002 |
| Female | 65+ | Cardiovascular diseases | Diabetes | 1.03 | <0.001 |
| Female | 65+ | Cardiovascular diseases | Musculoskeletal disorders | 0.72 | <0.001 |
| Female | 65+ | Cardiovascular diseases | Respiratory infections | 1.25 | <0.001 |
| Female | 65+ | Musculoskeletal disorders | Asthma/ COPD | 0.60 | 0.002 |
| Female | 65+ | Musculoskeletal disorders | Diabetes | 0.65 | 0.002 |
| Female | 65+ | Musculoskeletal disorders | Mood and anxiety disorders | 0.58 | <0.001 |
| Male | 65+ | Cardiovascular diseases | Asthma/ COPD | 0.96 | <0.001 |
| Male | 65+ | Cardiovascular diseases | Cancer | 1.66 | <0.001 |
| Male | 65+ | Cardiovascular diseases | Diabetes | 1.19 | <0.001 |
| Male | 65+ | Cardiovascular diseases | Respiratory infections | 1.95 | <0.001 |
| Male | 65+ | Musculoskeletal disorders | Diabetes | 0.65 | 0.002 |

**Table B.1.** Health-condition combinations showing evidence of positive additive interaction and used to define the syndemic-based measure. RERI values greater than zero indicate positive additive interaction, meaning that the joint effect of both conditions exceeded the expected sum of their individual effects.

### Appendix C: Top 10 ICC values by domain

| Domain | Candidate variable | ICC |
| --- | --- | --- |
| Age composition | **Age 41–60 (%)** | **0.616** |
|  | Age 0–20 (%) | 0.581 |
|  | *Age 61–80 (%)* | *0.486* |
|  | Age 15–24 (%) | 0.481 |
|  | Age 21–40 (%) | 0.436 |
|  | Age >80 (%) | 0.417 |
|  | Age 45–64 (%) | 0.322 |
|  | Age 25–44 (%) | 0.268 |
|  | Age ≥65 (%) | 0.189 |
|  | Age 0–14 (%) | -0.072 |
| Income | *Health insurance payment arrears (%)* | *0.525* |
|  | **Low income (%)** | **0.514** |
|  | Persons in the national highest 20% income group (%) | 0.465 |
|  | Mean income per income recipient (€1,000) | 0.453 |
|  | Persons in statutory debt restructuring (WSNP) (%) | 0.448 |
|  | Mean income per resident (€1,000) | 0.436 |
|  | Mortgage debt (%) | 0.434 |
|  | Persons in the national lowest 40% income group (%) | 0.389 |
|  | Employees (%) | 0.277 |
| Education | High education, attained, derived via total population (%) | 0.526 |
|  | *Medium education, attended (%)* | *0.525* |
|  | **Low education, attained, derived via total population (%)** | **0.505** |
|  | High education, attained (%) | 0.458 |
|  | Medium education, attained (%) | 0.437 |
|  | Low education, attained (%) | 0.405 |
|  | Low education, attended (%) | 0.393 |
|  | High education, attended (%) | 0.372 |
|  | Medium education, attained, derived via total population (%) | 0.040 |
| Migration background | **Non-Western migration background (%)** | **0.685** |
|  | Surinamese background (%) | 0.674 |
|  | Native Dutch background (%) | 0.593 |
|  | Other background (%) | 0.593 |
|  | Moroccan background (%) | 0.581 |
|  | Non-Western migration background, derived via total population (%) | 0.580 |
|  | *Western migration background, derived via total population (%)* | *0.578* |
|  | Moroccan background (%) | 0.564 |
|  | Western migration background (%) | 0.527 |
|  | Surinamese background (%) | 0.501 |
| Housing | **Multi-family housing (%)** | **0.654** |
|  | Single-family housing (%) | 0.654 |
|  | Multi-person households (%) | 0.519 |
|  | *Owner-occupied housing (%)* | *0.498* |
|  | Rental housing (%) | 0.496 |
|  | Institutional household (%) | 0.475 |
|  | Mean assessed property value | 0.435 |
|  | Housing corporation-owned housing (%) | 0.418 |
|  | Housing built before 2000 (%) | 0.390 |
|  | Housing built after 2000 (%) | 0.390 |
| Lifestyle and social factors | **Meets physical activity guidelines (%)** | **0.590** |
|  | Overweight (%) | 0.544 |
|  | Severe overweight (%) | 0.489 |
|  | *Social isolation (%)* | *0.486* |
|  | Obesity (%) | 0.460 |
|  | Frailty among adults aged ≥65 (%) | 0.455 |
|  | ≥3 life events (%) | 0.445 |
|  | Sports participation (%) | 0.430 |
|  | Difficulty moving (%) | 0.415 |
|  | More than one residential move (%) | 0.328 |
| Facilities access | *Schools within 3 km (number)* | *0.597* |
|  | **Distance to large supermarket (km)** | **0.546** |
|  | Walkability index (500 m radius) | 0.504 |
|  | Distance to childcare (km) | 0.463 |
|  | Distance to general practice (km) | 0.366 |
|  | Distance to school (km) | 0.186 |
| Physical environment | **Public green space (%)** | **0.602** |
|  | *PM₁₀ concentration* | *0.543* |
|  | NO₂ concentration | 0.540 |
|  | PM₂.₅ concentration | 0.527 |
|  | Business-area neighbourhood type (%) | 0.511 |
|  | Elemental carbon concentration | 0.506 |
|  | Tree cover over water (%) | 0.453 |
|  | Tree cover in built-up area (%) | 0.447 |
|  | Water cover (%) | 0.443 |
|  | Historic city-centre neighbourhood type (%) | 0.432 |
| Social vulnerability | Youth care use (%) | 0.552 |
|  | **Welfare benefit use (%)** | **0.525** |
|  | *Victim support clients (%)* | *0.524* |
|  | Long-term care use (%) | 0.453 |
|  | Sickness benefit use (%) | 0.446 |
|  | Pension benefit use (%) | 0.436 |
|  | Disability benefit recipients (%) | 0.414 |
|  | Other social benefit use (%) | 0.367 |
|  | WMO use (%) | 0.344 |
|  | Social assistance benefit recipients (%) | 0.308 |

**Table C1.** Intraclass correlation coefficients (ICCs) for up to ten of the highest-ranking candidate contextual variables within each domain. Bold indicates the variable selected for the primary analysis and italics indicate the substitute variable used in sensitivity analyses. Variable selection was based on ICC in combination with theoretical relevance, interpretability, and comparability with the mobility data.

### Appendix D: Full stepwise OLS model selection for outgoing mobility distance

The primary adjustment set for the neighbourhood-level residual analyses was selected using a stepwise OLS procedure. Contextual domains were first ranked according to their explanatory power for outgoing weighted mean mobility distance in domain-specific models. Domains were then added cumulatively in this order, and changes in adjusted R² were used to assess whether each additional domain improved model fit. The full stepwise inclusion process is shown in Table D.1.

Because mobility was measured at the Mezuro level, the stepwise OLS procedure was used as a pragmatic variable-selection strategy to identify contextual domains that explained regional mobility patterns. The procedure was not intended as a causal model of mobility.

| Step | Included domains | Adjusted R² | Δ Adjusted R² | p-value | Decision |
| --- | --- | --- | --- | --- | --- |
| 1 | Lifestyle | 0.388 | — | — | Retained |
| 2 | Lifestyle + Education | 0.377 | -0.011 | 0.595 | Not retained |
| 3 | Lifestyle + Education + Ethnicity | 0.367 | -0.010 | 0.571 | Not retained |
| 4 | Lifestyle + Education + Ethnicity + Housing | 0.612 | +0.245 | <0.001 | Retained |
| 5 | Lifestyle + Education + Ethnicity + Housing + Environment | 0.659 | +0.047 | 0.018 | Retained |
| 6 | Lifestyle + Education + Ethnicity + Housing + Environment + Vulnerability | 0.713 | +0.054 | 0.085 | Not retained |
| 7 | Lifestyle + Education + Ethnicity + Housing + Environment + Vulnerability + Facilities | 0.704 | -0.009 | 0.958 | Not retained |
| 8 | Lifestyle + Education + Ethnicity + Housing + Environment + Vulnerability + Facilities + Income | 0.732 | +0.028 | 0.046 | Retained |
| 9 | Lifestyle + Education + Ethnicity + Housing + Environment + Vulnerability + Facilities + Income + Age | 0.740 | +0.008 | 0.174 | Not retained |

**Table D.1.** Full cumulative OLS model selection for outgoing weighted mean mobility distance.

The dependent variable was outgoing weighted mean mobility distance. Domains were added cumulatively based on their rank in domain-specific models. Δ Adjusted R² indicates the change in adjusted R² relative to the previous step. The final adjustment set used in the primary residual analyses consisted of lifestyle, housing, environment, and income. Education and ethnicity were not retained because their inclusion did not improve model fit in the cumulative model, despite showing explanatory power in domain-specific models. Vulnerability, facilities access, and age were not retained because they did not provide statistically or substantively meaningful improvement after the retained domains had been added.

### Appendix E: Diagnostic analyses of Western migration background and outgoing mobility

Western migration background emerged as a strong area-level correlate of mean outgoing mobility distance in the sensitivity analyses. Because both migration background and mobility were analysed at the Mezuro-region level, additional diagnostic analyses were performed to assess whether this association was robust, whether it was driven by a single region, whether it reflected mobility distance or mobility volume, and whether it could be explained by simple socioeconomic proxies. These analyses were considered diagnostic and exploratory and were not used as primary evidence for individual-level mobility behaviour.

#### E.1 Mezuro area level association with outgoing mobility distance

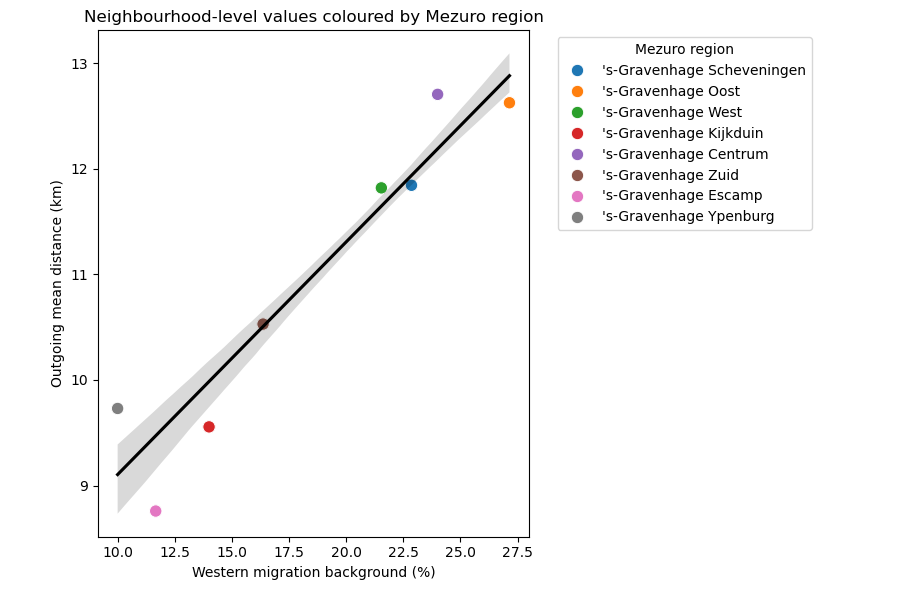
**Figure E.1.** Association between Western migration background and mean outgoing mobility distance across Mezuro regions.

The fitted line in Figure E.1 shows the linear association between the percentage of residents with a Western migration background and mean outgoing mobility distance. The association was strong at the Mezuro area level (ρ = 0.956; adjusted R² = 0.915), indicating that regions with a higher proportion of residents with a Western migration background had longer average outgoing mobility distances. This should be interpreted as an area-level association and not as evidence that individuals with a Western migration background travel farther.

#### E.2 Robustness to influential regions and population weighting

| Analysis | Correlation ρ | Adjusted R² |
| --- | --- | --- |
| Main Mezuro area level analysis | 0.956 | 0.915 |
| Population-weighted analysis | 0.957 | — |
| Leave-one-region-out minimum | 0.949 | 0.881 |
| Leave-one-region-out maximum | 0.977 | 0.944 |

**Table E.1.** Robustness of the association between Western migration background and outgoing mobility distance.

Leave-one-region-out values indicate the range of estimates obtained after excluding each Mezuro region one at a time. The population-weighted analysis weighted Mezuro regions by population size. Adjusted R² is not shown for the weighted correlation unless a corresponding weighted OLS model was estimated. Table E.1 shows that correlation and variance was not dominated by a single Mezuro region.

#### E.3 Mobility distance versus mobility volume

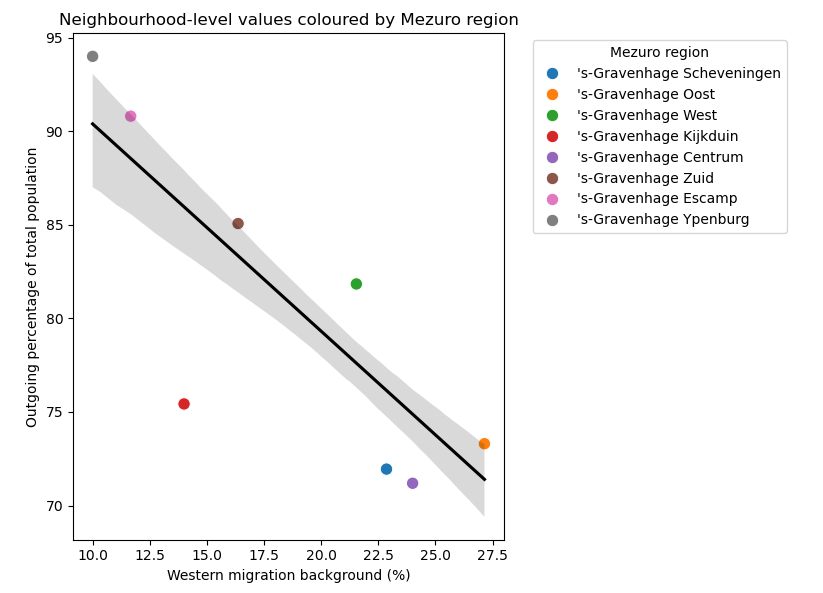
**Figure E.2.** Association between Western migration background and outgoing mobility ratio across Mezuro regions.

In Figure E.2, points again represent Mezuro regions. In contrast to the positive association with volume-weighted mean outgoing mobility distance, Western migration background was negatively associated with the outgoing mobility ratio. This indicates that the Western migration background signal relates primarily to the spatial reach of observed outgoing movements, rather than to a higher overall rate of outgoing movements. Because mean outgoing distance is itself weighted by movement volume across destinations, this finding should be interpreted as reflecting differences in the destination-distance profile of outgoing mobility, not as evidence of greater mobility volume.

| Mobility indicator | Correlation ρ |
| --- | --- |
| Mean outgoing mobility distance | 0.956 |
| Total outgoing movements | -0.162 |
| Outgoing mobility ratio | -0.793 |

**Table E.2.** Association of Western migration background with alternative mobility indicators.

These results in Table E.2 indicate that Western migration background was most strongly associated with the average distance of outgoing movements, but not with the total number or rate of outgoing movements. The negative association with the outgoing mobility ratio further suggests that this pattern should not be interpreted as higher overall mobility intensity. Rather, regions with a higher proportion of residents with a Western migration background appeared to have outgoing movements distributed toward more distant destinations.

#### E.4 Comparison of neighbourhood-level and Mezuro area level indicators

| Comparison | Neighbourhood level ρ | Mezuro area level ρ |
| --- | --- | --- |
| Neighbourhood-level vs postcode-derived Western migration background | 0.734 | 0.963 |
| Postcode-derived Western migration background vs outgoing mobility distance | - | 0.956 |
| Neighbourhood-level vs outgoing mobility distance | 0.714 | 0.953 |

**Table E.3.** Correlations between Western migration indicators and outgoing mobility before and after aggregation.

Table E.3 shows that the association was weaker at neighbourhood level, where mobility values were assigned from the corresponding Mezuro region. After aggregation to the Mezuro area level, the two Western migration indicators showed near-identical associations with mobility. This indicates that the strong association reflects a Mezuro area level compositional pattern rather than a coding artefact, while also highlighting that substantial within-region neighbourhood variation cannot be aligned precisely with mobility measured only at the Mezuro area level.

#### E.5 Relationship with socioeconomic and built-environment indicators

| Contextual indicator | Correlation ρ |
| --- | --- |
| Low income | -0.273 |
| WOZ value | 0.212 |
| Medium education | -0.648 |
| Meeting physical activity guidelines | 0.599 |
| Walkability | 0.473 |
| Public green space | -0.437 |

**Table E.4.** Correlations between Western migration background and selected contextual indicators.

Western migration background was not strongly correlated with simple socioeconomic proxies such as low income or WOZ value, as shown in Table E.4. Stronger associations were observed with education, physical activity guideline adherence, walkability, and public green space. This suggests that Western migration background may capture a broader regional composition pattern rather than acting only as a proxy for socioeconomic position.

#### E.6 Residual mobility–health associations after adjustment for Western migration background

| Outcome | Cross-scale neighbourhood ρ | Cluster-bootstrap CI |
| --- | --- | --- |
| Blood pressure medication | 0.02 | [-0.41, 0.44] |
| Diabetes medication | -0.22 | [-0.44, 0.06] |
| Cholesterol medication | -0.18 | [-0.46, 0.13] |
| Polypharmacy | -0.10 | [-0.36, 0.20] |
| Syndemic measure | -0.13 | [-0.44, 0.28] |

**Table E.5.** Residual mobility–health associations after Western migration adjustment.

Residual associations were calculated as Spearman correlations between residual mobility and residual health outcomes. Residuals were obtained by separately regressing outgoing mobility distance and each health outcome on Western migration background. These analyses, shown in Table E.5, address a diagnostic question: whether mobility–health associations remain after removing a single dominant Mezuro area level compositional correlate. They should therefore not be interpreted as primary adjusted associations or as replacing the main contextual-domain models.

Taken together, these diagnostic analyses indicate that Western migration background was a robust area-level marker of longer outgoing mobility distances. However, this pattern did not reflect higher mobility volume and should not be interpreted as an individual-level association. Instead, it shows that aggregated mobility indicators can capture regional population composition, spatial reach, and urban structure simultaneously. This supports the interpretation of outgoing mobility distance as a contextually and compositionally patterned measure rather than a standalone behavioural exposure.
